# Community action for newborn care and survival through participatory women’s groups and health workers in rural Bangladesh: a before-and-after implementation study of scale-up

**DOI:** 10.64898/2026.01.28.26344942

**Authors:** Edward Fottrell, Kohenour Akter, Abdul Kuddus, Sanjit Kumar Shaha, Badrun Nahar, Golam Azam, Tasmin Nahar, Anthony Costello, Kishwar Azad

## Abstract

**Background:** Community mobilisation through participatory women’s groups (PWGs) has been shown to be an effective intervention to improve maternal and neonatal survival in low-income settings, including Bangladesh. Despite WHO recommendations and scale-up in some contexts, the intervention has not been widely scaled-up in Bangladesh. To add to the existing evidence-base for PWGs and to renew calls for effective, scalable interventions to improve neonatal outcomes in the post-Sustainable Development Goals era, we report the design, implementation and evaluation of a volunteer-led model for PWGs delivered in rural Bangladesh in 2014/15.

**Methods:** Working in three rural unions in Faridpur, Bangladesh, we applied a volunteer-led, lower coverage and shorter duration PWG intervention. Mixed methods evaluation monitored key indicators of intervention delivery, uptake and receipt. Prospective quantitative surveys gathered data on birth outcomes, health care utilisation and essential newborn care practices. Data from before and after the implementation period were compared and interpreted in relation to historical trends in the study area and other rural areas of Bangladesh.

**Results:** 180 participatory women’s groups facilitated by 45 volunteer facilitators over a period of 15 months were successfully implemented giving a population coverage of one group per 500 population. An average of 32 (min.=18, max.=64) participants attended each PWG meeting, 42% of participants attended meetings on a monthly basis and 11% reported that they actively shared information from the PWGs with non-attenders. 30% of women of reproductive age and 54% of pregnant women participated in the. Focus group discussions with participants and community members revealed positive attitudes towards the groups. A change in trend in extended perinatal mortality rates was observed during the intervention period, corresponding temporally with indicators of improved rates of service utilisation and essential newborn care practices relative to the pre-implementation period.

**Conclusion:** The modified PWG intervention likely contributed to positive changes in delivery and neonatal care practices similar to previous studies in Bangladesh. The PWG model remains an important approach to community empowerment that could contribute to enhanced efforts to end preventable neonatal deaths as we move towards the end of the Sustainable Development Goal era and beyond.

## Background

Despite significant reductions in neonatal mortality globally, more than two million children die in the first 28 days after birth each year and reductions in neonatal mortality have slowed, with 64 countries likely to fall short of meeting Sustainable Development Goals (SDG) targets by 2030 unless urgent action is taken.^1^ This includes Bangladesh which is not expected to meet its SDG target on neonatal and child mortality this decade. Despite significant gains in maternal, neonatal and child health during the Millennium Development Goal and early Sustainable Development Goal eras, recent setbacks, cuts to international health funding, and decreased rates of neonatal mortality reduction demand new and accelerated efforts to scale-up implementation of effective interventions to end preventable neonatal deaths.^2-4^

Community mobilisation through participatory women’s groups (PWGs) practising Participatory Learning and Action (PLA) is one of two community approaches to improve neonatal survival recommended by WHO’s Global Strategy for Women’s Children’s and Adolescents’ Health.^5,6^ The PLA intervention is a process through which community members meet and, under the guidance of a lay facilitator, proceed through a four-phase cycle of identifying and prioritising threats to their health and neonatal survival, strategy development to address threats and maximise opportunities for positive health outcomes, implementation of strategies, and self-evaluation. A meta-analysis of seven trials evaluating the effects of PLA community mobilisation through PWGs found a 20% reduction in neonatal mortality (OR: 0.80; 95%:0.67-0.96).^7^ Coverage of such interventions is important, however. In Bangladesh, with coverage of approximately one group per 1400 population and participation of less than 3% of pregnant women, there was no evidence of effect on neonatal mortality (risk ratio 0.93 95% CI 0.80-1.09);^8^ when scaled to one group per 300 population and participation of pregnant women at more than 30%, mortality fell by 38% (risk ratio, 0.62 95% CI, 0.43-0.89).^9^

Despite good evidence and a WHO recommendation to implement PWGs in high mortality settings, there have been few attempts to scale-up the intervention to achieve maximum impact. In India, a large pragmatic effectiveness trial of PWG scale-up showed that government frontline workers and their supervisors could support PWG meetings in high-mortality settings in eastern India with significant impacts on neonatal mortality.^10^ There has been no large-scale implementation of PWGs in Bangladesh. Reasons for this may include the fact that the intervention is seen as time- and resource-intensive. To address this we designed, applied and evaluated a volunteer-led, budget-limited implementation of PWGs in rural Bangladesh to better understand factors that are essential to successful scale-up and sustainability, and those which may be modifiable. We reported our findings at community and stakeholder dissemination events in Bangladesh in 2015, and provide a summary here describing pragmatic implementation and evaluation processes and a call for renewed attention to community-led initiatives to realise the ambitions of global targets for child survival.

## Methods

### Study Setting

This implementation research study was a post-trial (trial registration ISRCTN01805825) intervention adaptation, scale-up and evaluation of PWGs. The study took place in three rural unions (the lowest administrative units in Bangladesh) in Sadarpur and Faridpur Sadar upazilas (sub districts) in Faridpur district, Bangladesh. Faridpur district has a population of over 1.7 million people in an area of just over 2000 square kilometres. The district has a mainly agricultural based economy, with the main crops being jute and rice. As in the rest of Bangladesh, primary care is provided at Union Health and Family Welfare Centres and at Community Clinics. In- and out-patient services are provided at sub-district (upazilla) health complexes and hospitals, and tertiary care is provided at district hospitals and medical college hospitals.^11^ Private service providers are also present in the area. Nevertheless, access to facilities and trained health care providers, short supply of medicines and low responsiveness of services remain challenges faced by the health care system in Faridpur district.

The three study unions were selected as they had previously been included as control clusters in our cluster randomised controlled trial of community mobilisation through PWGs and so were deemed accessible to the study team and benefited from being part of longitudinal surveillance systems of births and neonatal deaths since 2004^8,9^ – no new assignment occurred for the purposes of the current study. At the time of the study, the total population in the study areas was approximately 85,000 residing in approximately 20,000 households and representing approximately 16,500 women of reproductive age. Most of the population in the study areas is Muslim, with most of the remainder being Hindu.

### Original Intervention Review & Theory of Change

In PWGs, a local female facilitator supports groups in addressing health problems during pregnancy, delivery and neonatal periods using a four-phase Participatory Learning and Action (PLA) cycle. Through these four phases, women’s groups: (1) identify and prioritise problems in pregnancy, delivery and the neonatal period; (2) plan how to address these prioritised problems through locally feasible strategies; (3) implement their chosen strategies; and (4) evaluate their activities.^12^ The emphasis is on women identifying and solving their problems through their own efforts.

Implementation of PWGs during two cluster randomised controlled trials in Bangladesh consisted of salaried facilitators, who were local women of reproductive age with at least a high school qualification, being recruited, trained and paid to convene monthly women’s group meetings over a period of 20 months. Women’s groups were open to any members of the community, including men, although emphasis was placed on participation by women of reproductive age. Facilitators used visual aids (picture cards and flipcharts) to convey health messages and groups participated in discussions, roleplay, story-telling and wider community engagement to better prioritise the issues they faced and strategize their solutions to these. Group strategies varied between groups but three common strategies emerged: 1) voluntary emergency funds whereby women contributed money into a community fund for use in a health emergency; 2) wider awareness raising in the community whereby group members engaged with those outside of the group and held community meetings to share messages and learnings; and 3) transportation, whereby in certain areas, groups organised transport to local health and obstetric services.

Findings from the two cluster randomised trials in Bangladesh provided important lessons on the importance of intervention coverage. During the first trial, 162 women’s groups provided a population coverage of one group per 1414 population and, with low levels of participation among pregnant women and women of reproductive age, failed to achieve significant reductions in neonatal mortality.^8^ Subsequently, the second trial increased the number of groups to 810, giving a population coverage of approximately 1 per 300 population. Furthermore, concerted efforts were made by the implementing team and group facilitators to encourage pregnant women and women of reproductive age to participate in groups. Implementation at this level proved to be effective, leading to a 38% reduction in neonatal mortality.^9^

Based on a review of the previous trials in Bangladesh, as well as trials of PWG interventions in Nepal^13^, India ^14,15^ and Malawi^16,17^, we developed a framework of PWG intervention factors that we hypothesised to be essential for success and those we deemed modifiable (Figure 1).

**Figure 1:**
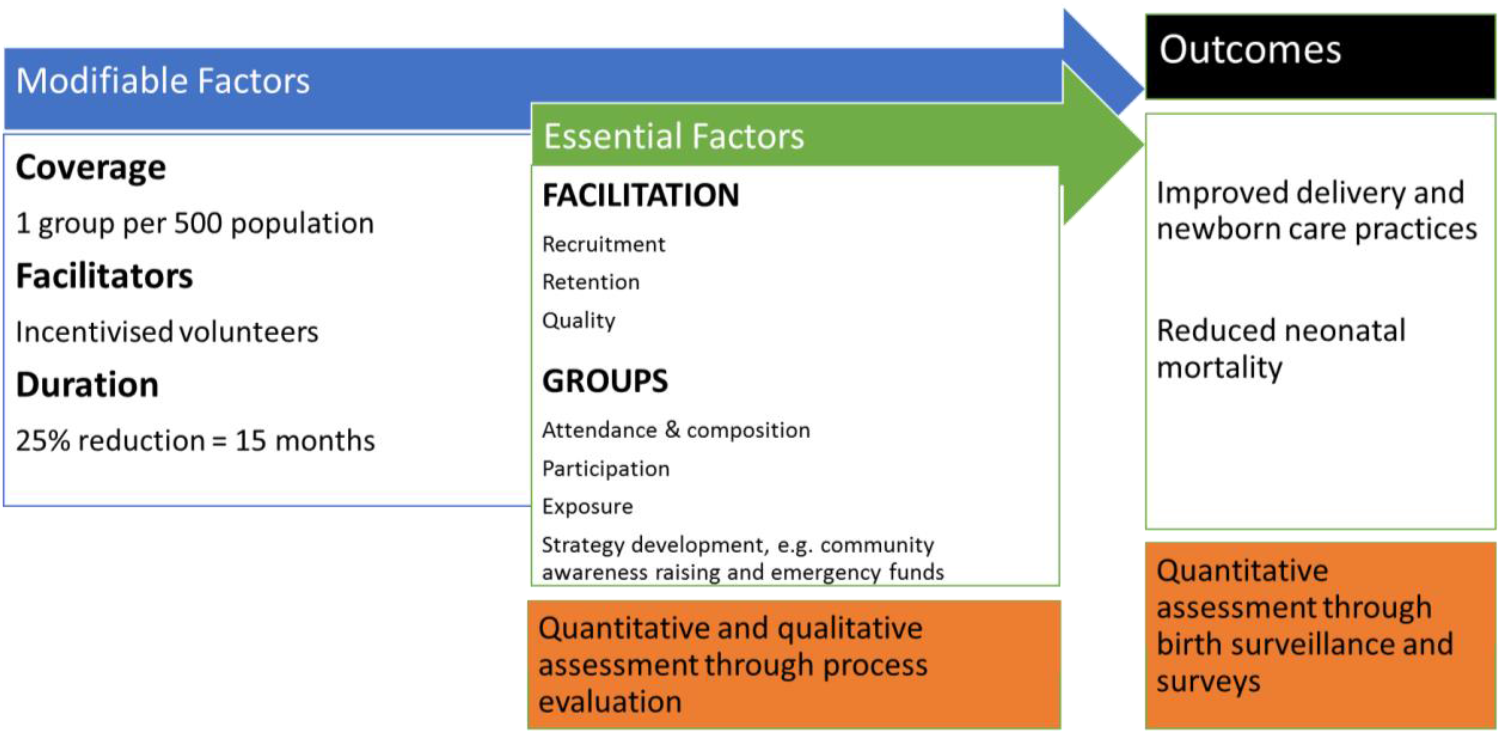
Framework of modifiable and essential factors and indicators of effective implementation of the PWG intervention. Red boxes represent project components and evaluation methods.

### Modifiable Factors

We hypothesised that three key features of the intervention were modifiable and could potentially increase the scalability, sustainability and cost-effectiveness of the intervention. These were, population coverage, the nature of group facilitation and intervention duration.

#### Coverage

As highlighted above, population coverage was an important determinant of intervention success in Bangladesh. Population coverage is considered critical for the key messages and lessons learned through PWGs to be disseminated beyond those who directly engage with the intervention. Achieving sufficient coverage of the key target population of women of reproductive age and pregnant women is crucial. Optimal population coverage is unknown, however, and the greater the number of groups the more the intervention costs. Coverage has varied across previous implementation of PWGs, being approximately 1 group per 756 in Nepal, 1 per 468 in rural India, 1 per 788 in urban India, and between 1 per 440 and 1 per 1200 in the two trials in Malawi.^13-17^ Findings from the meta-analysis of all trials indicated that with the participation of at least a third of pregnant women and with population coverage of 1 group per 450 to 750 population, PWGs were a cost-effective strategy to improve maternal and neonatal survival in resource-poor settings.^7^ Therefore, we decided to implement the intervention at a population coverage of 1 group per 500 population; this required 180 groups to be established.

#### Group facilitators

Previous implementation of PWGs in Bangladesh used salaried facilitators. However, our previous experience of 810 PWGs led by 45 facilitators (18 groups per facilitator per month) showed that, once established, women’s group members themselves often acquire the skills, commitment and enthusiasm needed to lead meetings on their own. Furthermore, implementation models elsewhere relied on volunteer or incentivised rather than salaried facilitators.^16^ Based on this, we expected that it would be possible to deliver the PWG intervention without paid facilitators. Instead, we decided to use a volunteer, cash-incentivised model whereby local women, low-level health staff and NGO workers would be recruited, given five days training through education and role-play on PLA methods and group facilitation techniques (and subsequent refresher training every six months) and motivated to form, hold and facilitate monthly PWG meetings. The role of the volunteer incentivised facilitators was to be the same as the salaried facilitators in our previous work, namely to convene monthly meetings and guide and supervise groups through the participatory learning and action cycle, with a cash honorarium being paid for each meeting conducted. We planned to recruit 45 volunteer facilitators, each responsible for running monthly PWG meetings in four locations (one per week), somewhat less than in our previous salaried model.

#### Intervention duration

Our previous experience of running monthly PWG meetings interventions over a period of 20 months indicated that this long duration might deter interest and participation in the groups, and thus effectiveness of the intervention overall. The intensity of PWG interventions in other settings has varied, from 10 monthly meetings in Nepal to 20 monthly in rural India, while in urban Mumbai groups met fortnightly over 18 months.^13-15^ Based on the process evaluation of our previous implementation in Bangladesh, we considered it feasible to proceed through the participatory learning and action cycle in a shorter timeframe and so revised and consolidated intervention content so that it could be delivered over 15 monthly meetings, i.e. a 25% reduction in duration.

### Essential Factors

The factors that we considered to be essential for the success of the PWG intervention can be broadly categorised as: facilitation – including recruitment, retention and quality of facilitators; groups – with emphasis on the attendance, composition, exposure; and participation – with the participatory nature of the groups and their strategies of raising community awareness and establishing local emergency funds being key.

#### Facilitation

The ability to recruit local women of reproductive age who resided in or within reasonable distance of PWG location, with sufficient education (set at 8 years of schooling in our project) and able to commit to undertake training and run PWGs on a regular basis is likely a key determinant of success. Further, retention of recruited facilitators is considered important to develop community trust and maintain a consistent approach to group facilitation. Finally, quality of group facilitation in terms of coordinating meetings, raising awareness of the intervention, recruiting participants to the groups, timeliness of meetings, use of intervention materials (facilitator manual, flipchart and picture cards) and ability to foster group participation are considered important determinants of success.

#### PWG attendance, composition & exposure

The success of any group-based intervention relies on individuals attending the groups and, often, consistent attendance across a series of meetings. In previous trials in Bangladesh, group size has been reasonably consistent at approximately 21 participants per group throughout the monthly meetings. However, community-wide impact of such interventions can only be achieved if intervention content and benefits are shared beyond those who directly participate. Process evaluation data from our implementation that resulted in significant neonatal mortality reductions shows that almost two-thirds of women residing in intervention clusters who did not directly participate in the intervention were, nonetheless, aware of it and approximately half reported that they had benefited from the intervention in some way. Further, a survey of participants showed that more than half of the women who attended groups actively discussed the intervention and its content with other, non-participants in their community. We therefore consider attendance in the PWGs and wider awareness of them to be crucial determinants of success.

A threshold attendance of women of reproductive age and pregnant women is considered particularly important to the effectiveness of the intervention as described elsewhere.^7,9^ Specifically, participation by 30% of pregnant women is considered an important criteria for effectiveness on neonatal mortality. Nevertheless, based on our experiences, we believe that wider participation, e.g. men and women outside the reproductive age range is also an important feature that promotes wider community mobilisation, dissemination of information and implementation of group-led strategies.

#### Participatory nature of group meetings

The level of active participation, interactive discussion and local ownership of groups, as opposed to passive, didactic learning is itself considered an essential feature of the PWG intervention. During previous implementation in Bangladesh this manifested as the selection and allocation of group attendees as ‘leaders’, who were women of reproductive age who took on a coordinating role of the group, ‘members’, women of reproductive age who volunteered for an active role in organising meetings and implementing strategies, and ‘participants’, who had a less active role but were nonetheless considered crucial to the community-wide acceptance, dissemination and broad mobilising nature of the intervention. Further, evidence of group-led implementation of strategies (phase 3 of the participatory learning and action cycle) are considered key indicators of participation essential for intervention success.

### Evaluation

Evaluation of our modified PWG intervention is based on two aspects. First, assessment of the extent to which elements deemed essential for intervention impact were achieved. This used process evaluation methods, combining a mix of quantitative and qualitative approaches to describe the recruitment retention and quality of facilitators, the number of PWGs formed, meetings held, PWG size and composition, community awareness of the intervention, and indicators of participation and wider community mobilisation. Second, direct measurement of community delivery and newborn care practices and neonatal mortality over the course of the project. This quantitative approach aimed to show trends in behavioural and mortality outcomes throughout the project and, when combined with the process indicators, helps to describe overall project success. We describe and compare process, behaviour and mortality indicators between the 2014/2015 implementation and the previous trial that showed intervention effectiveness in Bangladesh.

#### Process Evaluation

Data on group formation and facilitator recruitment and retention were obtained from project documentation. Quality of group facilitation was recorded by a PWG coordinator and three PWG supervisors, with each supervisor being responsible for 15 facilitators. The PWG coordinator documented every meeting that took place. The main responsibility of PWG supervisors was to observe facilitator performance, use of their facilitation skills and adherence to meeting plans during at least one PWG meeting per month. Each PWG facilitator maintained minutes and records of attendance, attendant characteristics (sex, age, marital status and pregnancy status, recorded using a structured form), strategies being implemented, and any important observations from each group meeting that they facilitated. These records were compiled by PWG supervisors and the coordinator every three months.

In 2015, we conducted a survey of a random sample of all group participants and facilitators using a structured questionnaire to gather data on their age, education and occupation. Based on a hypothetical parameter prevalence of 50%, 95% confidence, 5% error and a design effect of 2, we estimated that a sample size of 768 women would provide a representative sample of PWG participants. Adding 10% for non-response and rounding upwards our final target sample was 900 women and, assuming an average group membership of 25-27 participants, we randomly selected 34 facilitators and subsequently randomly selected one group per facilitator. All women in the selected groups were then included in our target sample and invited for interview.

We also conducted a total of 15 focus group discussions with women of reproductive age who did attend PWGs (n=6), women of reproductive age who did not attend PWGs (n=6), and PWG facilitators (n=3) to explore receipt of the intervention, satisfaction of participants with group facilitation, barriers to group participation, and challenges faced in group facilitation. Focus group discussions were conducted in Bangla with 6-12 respondents in each group and were led by a researcher with training in social science and qualitative research methods. All discussions were recorded by digital recorder and notes was also taken. The FGD recordings were transcribed in Bangla.

A total of 17 semi-structured interviews with PWG facilitators, supervisors and community stakeholders were also conducted to better understand perceptions of the intervention and challenges faced in implementation in this context. Interviews were conducted with local health workers (n=4), a Union Chairman (local leader) (n=1), PWG participants (n=2), PWG facilitators (n=3), the PWG coordinator (n=1), the PWG supervisors (n=3), and government officials (n=3). Participants were selected purposively in relation to their knowledge and association with the project and data were collected to saturation.

Simple quantitative analysis is used to describe the characteristics of participants in PWGs and the volunteer facilitators, as well as those who do not actively engage with the intervention. Thematic analysis of focus-group data and qualitative interviews with key stakeholders was used to further elucidate the receipt and perceptions of the intervention. In the current analysis, we only present broad findings that relate to the essential factors of the intervention as previously described.

#### Quantitative mortality and behaviour indicators

From April 2013 to June 2015 (Periods C and D in figure 2) we used a low-cost prospective surveillance system to record all births and their outcomes (live birth, stillbirth, neonatal death) and all deaths during pregnancy and up to 6 weeks post-partum in the three study unions. Based on a previously described surveillance system^18^, each of 82 key informants, all traditional birth attendants, were responsible for identifying all women that give birth and all deaths to women of reproductive age within a geographical area of around 200 households. At the time of the study, TBAs conducted the majority of deliveries in the study area and were therefore in a good position to identify births, irrespective of whether they attended them. Each TBA was visited every two weeks by a salaried monitor who collected identifying details of all events registered by the TBAs. The monitor subsequently verified all events through household visits, and, for deaths to women of reproductive age, establish whether the death occurred during pregnancy or up to six weeks postpartum. TBAs received a Tk100 ($1 approx. in 2014) honorarium for each accurate notification. This process of birth and death surveillance provided the essential information to measure trends in mortality rates (total, early neonatal mortality and late neonatal mortality), stillbirth rate, perinatal mortality rate and pregnancy-related mortality rate throughout the period of implementation.

**Figure 2:**
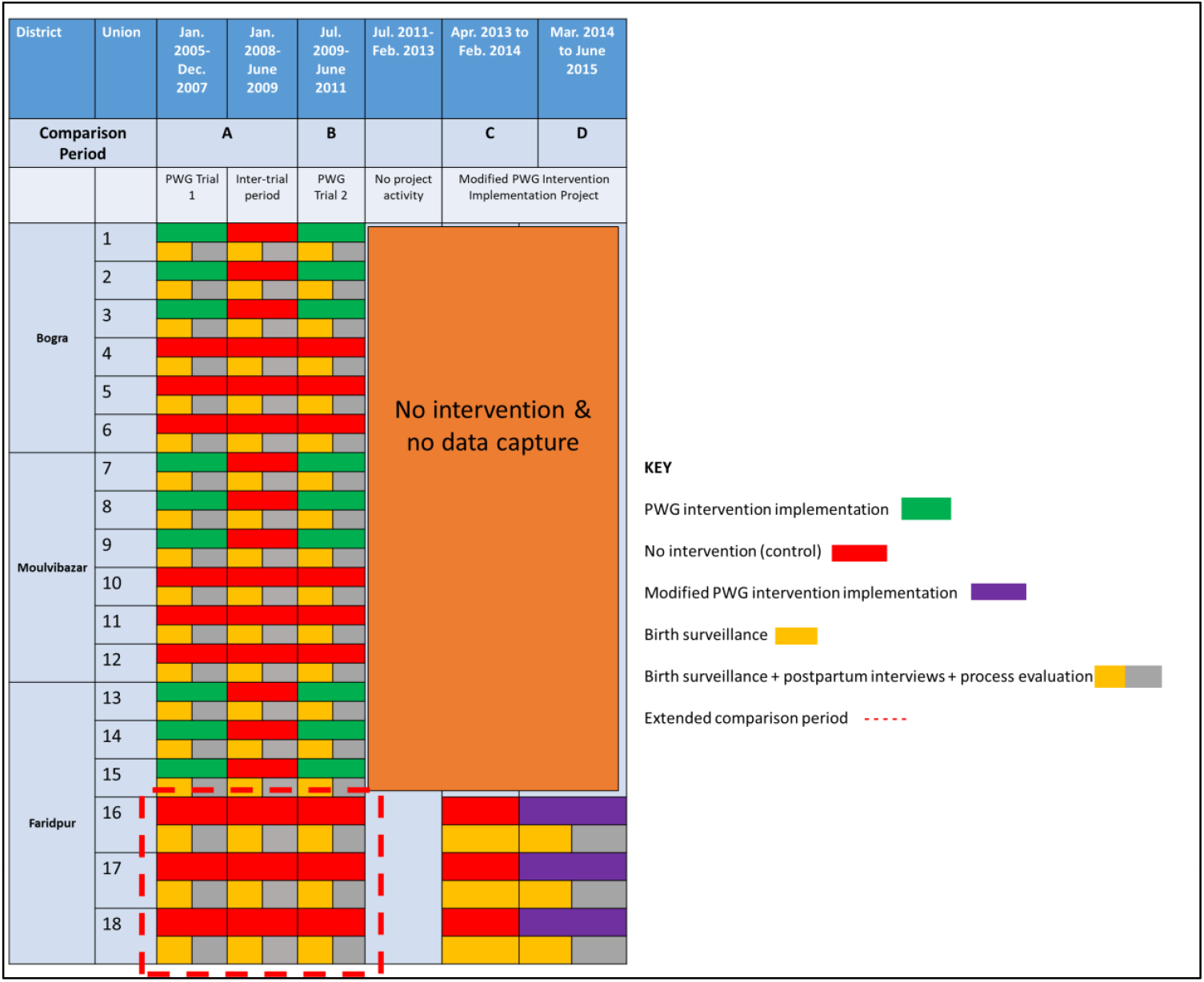
Data sources for implementation evaluation. Process evaluation uses and compares data gathered throughout Period D, the time of the modified PWG intervention implementation, and Period B, the 2009-2011 PWG trial period. Mortality indicators relate to Periods A, B, C and D. Newborn care practices and health service utilisation data are compared between periods A, B and D. Period A data are only used from the three Faridpur study unions to provide and extended comparison period in the study areas. ^*^*Note that a period of no intervention during 2007 and 2008 is not shown in the figure for simplicity*.

Throughout the period of modified intervention implementation (March 2014 to June 2015, Period D Figure 2), monitors re-visited households between 6 to 52 weeks following the registered event to interview the mother (or in the case of a maternal death, the closest family member) and gather data on delivery and newborn care practices using a brief structured questionnaire, adapted from our previous trials. Process indicators of awareness of the PWG intervention and participation in it were also gathered. Details of data collection and intended use of the data was explained to respondents in their homes and opportunities were given for clarification or discussion of issues before the mother decided whether or not to be interviewed.

#### Comparison to previous trial data

A key aspect of the analysis and interpretation of evaluation data is comparison of process indicators, outputs and outcomes between the modified, budget limited approach and the higher-cost, proven implementation documented as part of our trial between 2008 and 2011 (Figure 2 Period B). Comparisons are made between the modified implementation period (Period D) with the overall trial intervention and control clusters (Period B) and, separately, the specific three current study unions immediately prior to intervention implementation (Period C) and over an extended period of surveillance from 2005 to 2011 (Figure 2 Periods A and B for Faridpur control clusters only, indicated by the dotted outline). All quantitative analysis of behavioural and mortality indicators are based on cluster level summary measures, considering each Union as a separate cluster. Rather than using formal statistical testing between diverse datasets, comparisons rely on temporal observations and interpretations based on previous evaluations and discussions of PWG interventions.

### Ethics

Project activities were reviewed and approved by the Ethical Review Committee of the Diabetic Association of Bangladesh (BADAS-ERC/EC/13/00106).

## Results

### Process evaluation

45 women of reproductive age were recruited as facilitators in January 2014. The recruitment process took longer than expected due to difficulties finding women with the requisite education who were not already employed elsewhere. Education level of facilitators was lower in the modified implementation than in our previous implementation, with a requirement of completion of 8^th^ grade education in the current implementation compared to higher-secondary education (two years post-secondary education) previously. Facilitators received five days training in facilitation techniques, community mobilisation, participatory and communication skills and the use of visual aids. Eighteen of the 45 recruited facilitators (40%) subsequently resigned from their positions and were replaced. This is a similar level of retention as was observed when we used salaried facilitators in our previous trial in Bangladesh (36%). Reasons for resignation included prioritisation of study for impending exams (n=4), uptake of better paid employment (n=2), pregnancy (n=2), and family reasons (n=10), which included domestic obligations, dissatisfaction with honorarium payment and disapproval of the job by family members.

180 PWGs were established within an estimated study population of 89,000, giving a population coverage of one PWG per 494 population. Monthly meetings were held from March 2014 to June 2015. All groups completed the full programme of 15 meetings, although some meetings were delayed due to heavy rains, the harvesting season and religious festivals, including Ramadan. Eight hundred and eighty-four women out of a target 900 participated in our random sample survey of group participants’ socio-demographic characteristics, representing a response rate of over 98%. The mean age of PWG participants was 29 years, 88% were Muslim, 84% were literate, 85% had completed at least primary education and 97% were currently married. An average of 32 (min.=18, max.=64) participants attended each PWG meeting, 42% of participants attended meetings on a monthly basis and 11% reported that they actively shared information from the PWGs with non-attenders.

To estimate the proportion of women of reproductive age that participate in the PWGs, we divided the number of women of reproductive age who attended PWG meetings (derived from facilitator registers, n=4960) by the estimated population of women of reproductive age in the study areas (16500), i.e. 30%. Of all women interviewed as part of the prospective birth surveillance system, 75% reported awareness of the PWG intervention and 54% reported participation. Table 1 shows process indicators and comparison to the same indicators from our previous trial implementation of PWGs (i.e. comparison between periods D and B in figure 2). Our modified implementation of PWGs achieved higher overall attendance and higher rates of attendance by women of reproductive age and pregnant women, but lower dosage, lower levels of awareness of the intervention among women who were pregnant and lower levels of dissemination of information overall.

**Table 1:**
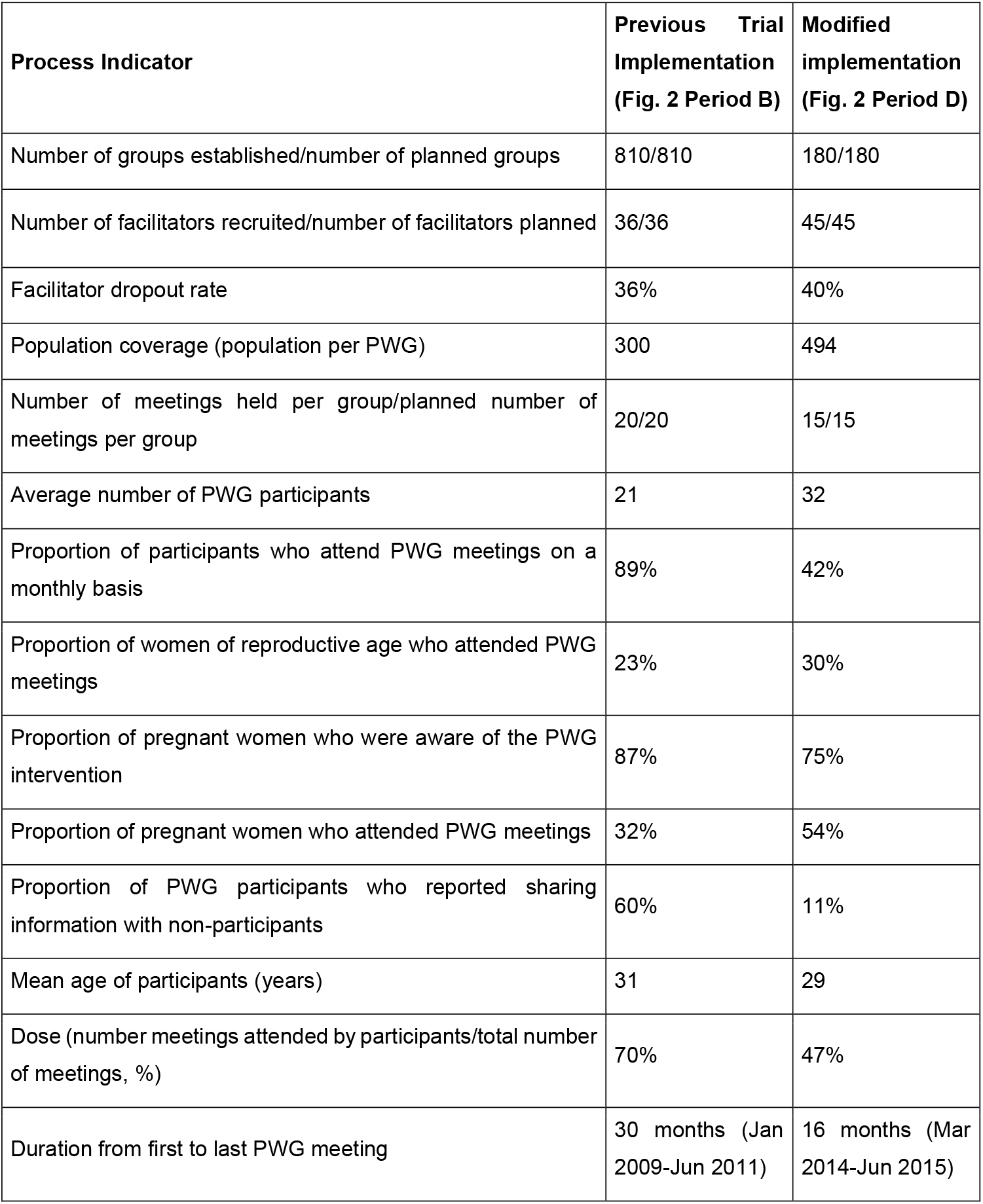
Quantitative process indicators from the budget- and time-limited implementation in three unions and from previous trial implementation in 9 unions.

| **Process Indicator** | **Previous Trial Implementation (Fig. 2 Period B)** | **Modified implementation (Fig. 2 Period D)** |
| --- | --- | --- |
| Number of groups established/number of planned groups | 810/810 | 180/180 |
| Number of facilitators recruited/number of facilitators planned | 36/36 | 45/45 |
| Facilitator dropout rate | 36% | 40% |
| Population coverage (population per PWG) | 300 | 494 |
| Number of meetings held per group/planned number of meetings per group | 20/20 | 15/15 |
| Average number of PWG participants | 21 | 32 |
| Proportion of participants who attend PWG meetings on a monthly basis | 89% | 42% |
| Proportion of women of reproductive age who attended PWG meetings | 23% | 30% |
| Proportion of pregnant women who were aware of the PWG intervention | 87% | 75% |
| Proportion of pregnant women who attended PWG meetings | 32% | 54% |
| Proportion of PWG participants who reported sharing information with non-participants | 60% | 11% |
| Mean age of participants (years) | 31 | 29 |
| Dose (number meetings attended by participants/total number of meetings, %) | 70% | 47% |
| Duration from first to last PWG meeting | 30 months (Jan 2009-Jun 2011) | 16 months (Mar 2014-Jun 2015) |

Several PWG strategies emerged from the intervention. The most popular strategies were the creation of group emergency funds and wider community awareness raising. Emergency funds were created by all groups, providing a source of financial help to women in times of need and without the need for collateral. Reasons for allocating funds to women included serious illness of the woman or her baby or any medical emergency within the village. Financial support was usually considered to be a loan which the beneficiary would pay back to the group. Some groups who initially adopted this strategy later abandoned it due to issues of managing and disbursing the funds and instead opted for different strategies, such as establishing systems to arrange transport to health facilities.

A total of 47 community meetings held by the PWGs throughout the project period. The purpose of these community meetings was to engage the wider community and raise awareness of the prioritised problems and the barriers that women face during pregnancy and childbirth. The community meetings also gave PWG participants opportunities to discuss their strategies to overcome the problems and barriers they face with the wider community. Anecdotal evidence and case studies of the consequences of this wider community engagement were obtained by the Process Evaluation Manager and include one scenario where the Union Parishad Chairman, influenced by the PWGs, advocated for and ultimately succeeded in creating a paved path to one village to enable access by motorised vehicles, e.g. ambulances.

Findings from the FGDs revealed positive attitudes towards the intervention. PWG participants described effects of the intervention increasing knowledge and awareness of health issues affecting pregnant women, mothers and their babies. They stated that the knowledge they gained from the meetings was relevant to them and comprehensible and reported that they felt confident sharing the information with others outside the group.

> ‘*We can learn many things from the meeting. Previously I did not take calcium or iron tablets; neither did I take any rest. However, after attending these meetings I understand the importance of taking calcium and iron tablets and resting for two hours. Prior to these meetings I did not realise that the weakness I was feeling may have been due to anaemia. I also learnt which types of food I should take regularly, which are nutritious, cheap, and which could be grown in the homestead*.*’* (FGD, Woman of Reproductive Age, PWG Attendee)

Data from women who did not attend PWG meetings highlighted barriers to participation, which included distance to meeting place, the perceived time commitment needed, and competing responsibilities, such as domestic work responsibilities. Some respondents stated that they did not attend as there was no material incentive for doing so. In some cases, respondents reported that their husbands or mothers-in-law or other influential elders prevented them from attending, citing perceived risks of being influenced by individuals or groups outside the family. A small number of young women reported that they were unable to participate due to ‘purdah’ and/or fear of ‘evil eye’ whereby, newly married or newly pregnant women are restricted from going out of the home for a period of time. Nevertheless, one FGD participant who had previously not been allowed to attend groups described how attitudes had changed as the community became more familiar with the intervention:

> ‘*A lot of positive changes have taken place due to these women’s groups meetings. For example, my mother-in-law now tells me to attend the meeting first and do household work later*.’ (FGD, Woman of Reproductive Age, PWG Attendee)

Interviews with PWG participants and facilitators indicated that meetings were conducted well and facilitators were seen to be highly motivated by themselves and PWG participants. Nevertheless, facilitators did state that they wanted greater incentives for running groups and some felt that the intervention materials (e.g. the flipchart and picture cards) would be better as posters or in a digital format.

### Behaviour and mortality indicators

A total of 2030 births were registered in the study unions during the 11 months (April 2013 to February 2014) prior to intervention implementation (figure 2, period C). Of these, 1685 (83.0%) were to women who had permanently resided within the study unions and were eligible for inclusion in our study. Neonatal mortality rate during this pre-implementation period was 21.8 per 1000 livebirths and stillbirth rate was 17.7 per 1000 births (Table 2). A further 3466 births were registered during the 16 months of project implementation (March 2014 – June 2015, figure 2 period D), 2286 of which were to permanent residents (66.0%). Overall neonatal and stillbirth mortality rates for the implementation period were similar to the pre-implementation period, being 20.4 per 1000 livebirths and 17.9 per 1000 births, respectively.

**Table 2:** Births deaths and mortality rates by study period and in comparison to the previous PWG trial.

|  | Trial Period July 2009-June 2011<br>(Fig 2. Period B) |  | Faridpur<br>(3 unions) |  |  |
| --- | --- | --- | --- | --- | --- |
|  | Control* | Intervention | 2005-2011<br>(Fig 2. Periods A & B, control clusters only, no intervention) | April 2013-February 2014<br>(Fig 2. Period C, no intervention) | Mar 2014-Jun 2015<br>(Fig. 2 Period D, modified intervention) |
| Total births | 8834 | 9106 | 13243 | 1685 | 2286 |
| Live births | 8602 | 8819 | 12829 | 1655 | 2244 |
| Stillbirths | 232 | 287 | 414 | 30 | 42 |
| Early neonatal death | 210 | 148 | 336 | 30 | 39 |
| Late neonatal deaths | 61 | 39 | 84 | 7 | 6 |
| Total neonatal deaths | 271 | 187 | 420 | 37 | 45 |
| Perinatal deaths | 442 | 435 | 750 | 60 | 81 |
| Pregnancy-related deaths | 20 | 13 | 31 | 3 | 4 |
| *Cluster mean rates* |  |  |  |  |  |
| Stillbirth mortality rate per 1000 births | 26.0 | 31.6 | 31.4 | 17.7 | 17.9 |
| Neonatal mortality rate per 1000 livebirths | 30.1 | 21.3 | 32.0 | 21.8 | 20.4 |
| Perinatal mortality rate per 1000 births | 48.9 | 47.9 | 56.2 | 34.8 | 35.1 |
| Pregnancy-related mortality ratio per 100000 livebirths | 276.1 | 153.4 | 229.4 | 166.2 | 176.9 |

\*Control clusters excluding tea-garden residents as detailed in <sup>9</sup>

Using all previously available data from the three study unions in Faridpur (figure 2, periods A and B for Faridpur control clusters only), extended perinatal mortality trends are shown as moving 6-month averages in Figure 3. Despite a data gap from mid-2011 to April 2014, an overall pattern of falling mortality is observed over time. Furthermore, trends during the immediate pre-intervention period and during the intervention are in opposite directions, with apparent declining mortality trends during implementation.

**Figure 3:**
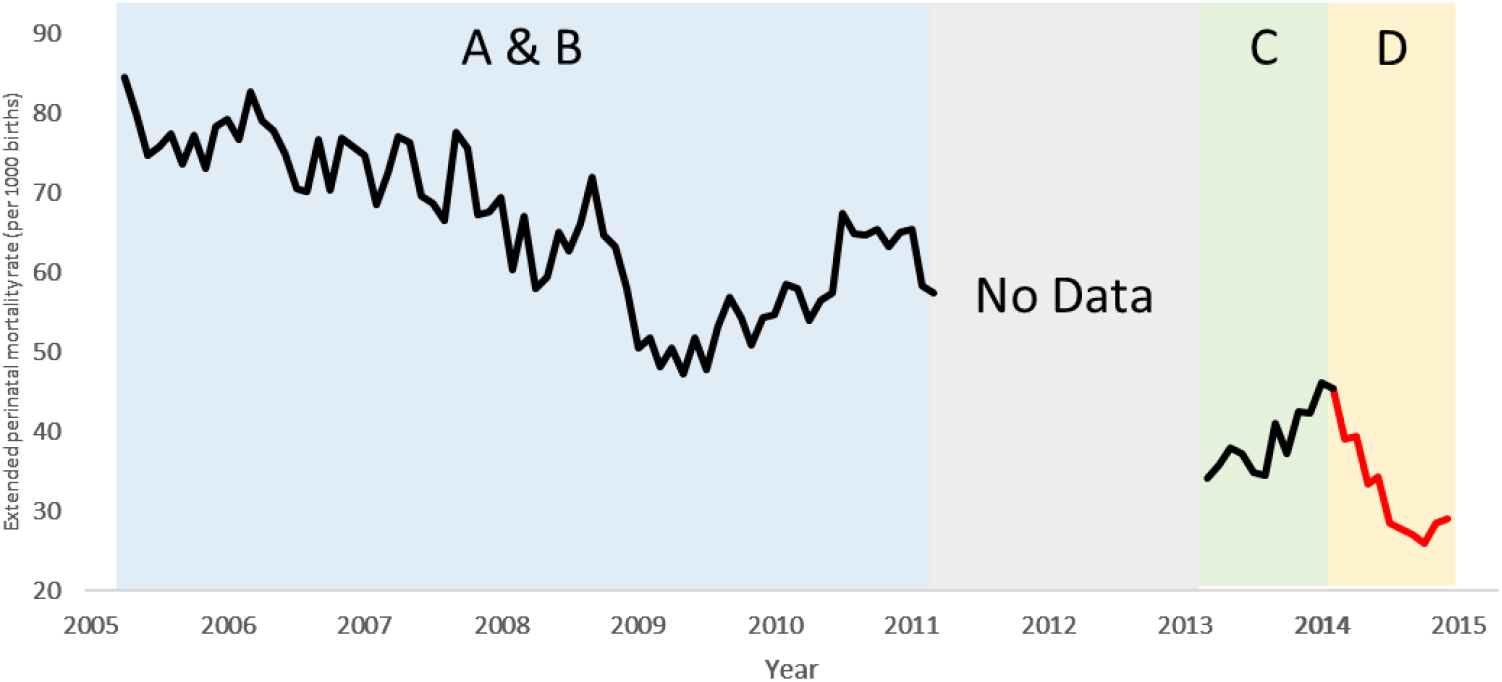
Moving six-month average perinatal mortality rate in the Faridpur study areas from 2005-2011 (Periods A & B) and before (C) and during (D) implementation of the PWG intervention.

Of the 2286 births registered during the implementation period and eligible for inclusion in our analysis, postpartum interviews were completed for 2134, representing a response rate of 93.4%. There were no significant differences in mortality outcomes between responders and non-responders to the survey. Quantitative data on birth practices, home-based newborn care, thermal care, breastfeeding, and health service utilisation are shown in Table 3. Facility deliveries and the number of births attended by a medical professional increased during the project implementation period relative to previous years, from 33.1% to 54.9% (facility deliveries) and 36.6% to 61.4% (medical birth attendant). Other care seeking practices during the antenatal and postnatal/postpartum period also increased considerably.

**Table 3:** Home-based newborn care, thermal care, breastfeeding, and health service utilisation by study period and in comparison to the previous PWG trial.

|  | Trial Period 2008-2011, 9 unions (Fig 2. Period B) |  | Faridpur, 3 unions |  |
| --- | --- | --- | --- | --- |
|  | Control* | Intervention | 2005-2011 (Fig 2. Period A & B, control clusters only, no intervention) | Mar 2014-Jun 2015 (Fig. 2 Period D, modified intervention) |
|  | % | % | % | % |
| **HEALTH SERVICE UTILISATION** |  |  |  |  |
| *Antepartum* |  |  |  |  |
| 4 or more ANC checkups by medical provider | 14.8 | 18.9 | 14.9 | 31.4 |
| *Intrapartum* |  |  |  |  |
| Facility deliveries | 27.7 | 26.8 | 33.1 | 54.9 |
| Birth attended by a medical provider<sup>c</sup> | 30.8 | 28.4 | 36.6 | 61.4 |
| *Postpartum* |  |  |  |  |
| Mother received check-up in first 6 weeks by a formal provider<sup>d</sup> | 23.1 | 22.5 | 25.8 | 45.1 |
| Infant received check-up in first 6 weeks by a formal provider<sup>b</sup> | 23.6 | 20.4 | 25.0 | 52.9 |
| **HOME DELIVERY PRACTICES** |  |  |  |  |
| Birth attendant washed hands with soap<sup>a</sup> | 83.8 | 91.3 | 70.5 | 71.5 |
| Birth attendant used safe delivery kit<sup>a</sup> | 15.5 | 29.1 | 9.3 | 26.0 |
| Birth attendant used plastic sheet<sup>a</sup> | 62.1 | 72.5 | 52.0 | 77.6 |
| Birth attendant used boiled or new instrument<sup>a,b</sup> | 98.9 | 99.5 | 99.0 | 93.3 |
| Birth attendant boiled thread<sup>a,b</sup> | 56.2 | 66.8 | 58.5 | 70.9 |
| Cord left undressed or dressed with antiseptic<sup>a,b</sup> | 25.4 | 36.9 | 11.1 | 73.2 |
| **THERMAL CARE OF NEWBORNS (HOME DELIVERIES)** |  |  |  |  |
| Infant wiped and wrapped within 10 mins of delivery<sup>a,b</sup> | 33.3 | 36.0 | 34.7 | 15.9 |
| Infant not bathed in first 24 hours<sup>a,b</sup> | 70.1 | 90.0 | 74.7 | 87.2 |
| Infant placed on mother's skin within 30 mins of delivery<sup>a,b</sup> | 51.7 | 80.7 | 70.3 | 71.3 |
| **EARLY INFANT FEEDING PRACTICES (ALL LIVE BIRTHS)** |  |  |  |  |
| Exclusive breastfeeding for at least 6 wks<sup>b</sup> | 74.4 | 77.9 | 60.0 | 81.7 |

a - home births only; b – livebirths only \*Control clusters excluding tea-garden residents as detailed in <sup>9</sup>

For home births, the majority of indicators show higher levels of essential newborn care practices during the project implementation compared to pre-intervention levels, for example use of Safe Delivery Kits, delivery on plastic sheets, and hygienic cord care. Furthermore, for several indicators the levels of such behaviours during the project implementation increase by similar or larger amounts and to levels similar to those achieved in intervention clusters in the previous PWG trial. Smaller changes are observed in the proportion of mothers who reported skin to skin care within 30 minutes of delivery during project.

The proportion of women reporting exclusive breastfeeding for at least 6 weeks was somewhat lower in the Faridpur control areas in 2005-2011 than even the control areas of the full PWG trial. However, it increases to 81.7% during the modified intervention implementation period.

## Discussion

In 2014 and 2015, we adapted and applied a lower-coverage, volunteer-led implementation of the PWG intervention over a period of 15 months in three unions of Faridpur district. Mixed method evaluation of the implementation process, receipt of the intervention, and indicators of behaviour change relating to health care utilisation, home delivery practices, essential newborn care, thermal care and breastfeeding suggest that the intervention was successfully delivered and is likely to have contributed to positive changes in delivery and neonatal care practices. Mortality rates immediately before and during intervention implementation were very similar, but trend data indicate opposite directions, with apparent falling mortality during the intervention period.

Despite reporting activities and data from a decade ago, our experiences and findings remain highly relevant in a context of stagnating progress and missed goals in the late SDG era. Our study was based on a simple conceptualisation of essential and modifiable elements of the PWG intervention and provides a framework with which to implement and evaluate the intervention. Though likely to be an oversimplification of the intervention, our framework (figure 1) is practical and aligned with a mixed methods process evaluation approach that utilises existing and new data to better understand potential mechanisms of intervention effect.^19^ We have described specific intervention adaptations and, in terms of implementation research, our outcome measures of PWG delivery, reach and coverage, as well as behaviours related to delivery and newborn care practices can each be considered critical ends in and of themselves. Combined, these measures shed light on how the adapted PWG intervention was implemented and received and the mechanisms that plausibly contribute to effectiveness in terms of mortality reduction. The ability to compare our study data with outcomes measured both before intervention delivery in the study population and from previous implementation in similar populations enhances our ability to discuss observed changes in relation to possible intervention effects although, without a contemporary comparison area, interpretation of cause and effect requires caution.

Our coverage, facilitation and duration modifications of the PWG approach to community empowerment are likely to have affected the overall level of exposure or ‘dose ‘of intervention within the community. Despite this, we observed improved delivery practices, care-seeking and preventative care practices that have been identified previously as important PWG contributors to improved newborn survival ^20,21^. Observed behavioural outcome measures and comparisons to historical data indicate positive changes that echo those observed in the previous cluster randomised trial of the intervention in Bangladesh and, for some indicators, exceed the magnitude of positive change one might expect based on previous work. For example, increased facility deliveries and birth attendance by medical professionals is a change not observed in the earlier trial of women’s groups in Bangladesh or in other settings where women’s groups have been evaluated. Indeed, the smaller change in the proportion of mothers who reported skin to skin care within 30 mins of delivery during project might reflect changes in who actually delivered the baby, with a greater number of births being attended by medical professionals during the project period, where procedures and routine practices might be a barrier to rapid skin to skin contact. These changes are likely to reflect wider, secular improvements in service utilisation in Bangladesh during the implementation period. For example, we are aware of other non-governmental organisation initiatives providing home visits, health worker training, and antenatal care in Faridpur during this time. Notably, however, implementation of the Government of Bangladesh’s Maternal Health

Voucher Scheme to reduce barriers to maternal, newborn and child health service utilisation^22-25^ had not yet started in the study areas when we conducted our study (Faridpur Civil Surgeon – personal communication, 2026). Similarly, changes in perinatal mortality, essential newborn care, home delivery practices and breastfeeding may also reflect random fluctuations and secular trends that our study design cannot account for.

Implementation research is sometimes discussed in terms of a trade-off between rigour and relevance, with relevance occasionally being seen as less rigorous due to the lack of direct counterfactual comparisons and the advantages these offer. Conversely, the greater the rigour of an evaluation, e.g. through randomisation of the intervention and comparison to a control, the more artificial and less directly informative about scalability and impact in real-world settings the findings can be.^26^ The effectiveness of PWGs has been demonstrated in cluster RCTs so, in an attempt to progress from proof of effectiveness to intervention use on a larger scale we prospectively embedded evaluation and measurement of process and outcome indicators into the process of intervention implementation. The evidence from this implementation project therefore requires interpretation in relation to what we already know about the effectiveness of PWGs, the sorts of impacts expected and the plausibility of observed changed being attributable, at least to some extent, to the intervention.

The Global Strategy for Women’s, Children’s and Adolescents’ Health (2016-2030) is a roadmap for ending preventable deaths (survive), ensuring health and well-being (thrive) and expanding enabling environments (transform).^27^ Community empowerment was identified as a priority for the transformative component of this agenda.^28^ In mid-2025 the Partnership for Maternal, Newborn & Child Health (PMNCH) launched a new 2026-2030 strategy as a successor platform to help operationalise the global strategy goals, with renewed emphasis on community-centred and led action and scale-up of proven, cost-effective interventions.^29^ Combined with trial findings from scale up initiatives led by Accredited Social Health Activists (ASHAs) and their supervisors in rural India, findings from our study contribute to an timely understanding of how the PWG approach to community empowerment might continue to be effective when brought to scale.^10,30^ Realising this will require government buy-in and recognition of the value of PWGs.

Our findings also contribute to understanding the causal pathways behind the success or failure of PWG interventions. We hypothesised, *a priori*, that certain elements were essential to intervention success, including the recruitment, retention and high quality of group facilitators, adequate attendance of women of reproductive age and pregnant women at group meetings, and strategies to raise awareness of the intervention even among those not directly participating (e.g. through wider community engagement). Our process data suggest these elements were achieved in our scaled-up implementation. Further, we observed improvements in behaviours that were also seen in previous PWG studies^31^ and that plausibly contribute to the improved neonatal survival. Other mechanisms through which PWGs may work are also likely to exist and were not measured in our study, however. Socio-cultural norms, the physical environment and the specific foci of PWG discussions and strategies are likely to differ between groups and investigation of these in future implementation may help to describe the intervention process, exposure experiences and mechanisms of action to a greater degree.

Despite progress in child mortality overall, neonatal mortality rates in Bangladesh today remain high, with the greatest burdens in poorer populations and rural communities.^32,33^ As we near the end of the SDG-era, it is timely to reflect on progress and opportunities to address the structural, health-system and socioeconomic determinants of neonatal survival. Revisiting the 2014 WHO recommendation for the implementation of community mobilisation through facilitated PWGs,^6^ our findings are a reminder of an effective, equitable,^34^ scalable, and potentially sustainable approach that may be adapted to current demographic and epidemiological contexts.

## Data Availability

Data available upon request to the authors.

## Acknowledgements

This project was funded by a grant from the WHO Implementation Research Platform awarded to the Diabetic Association of Bangladesh. We are grateful to Dr Nhan Tran for input to project design and throughout the project. We thank Dr Ashit Ranjan Das, Civil Surgeon of Faridpur District for his support.

## Notes

### Competing Interest Statement

The authors have declared no competing interest.

### Clinical Trial

ISRCTN01805825

### Funding Statement

This study was funded by WHO Implementation Research Platform.

### Author Declarations

The ethics review committee of the Diabetic Association of Bangladesh gave ethical clearance for this work (BADAS-ERC/EC/13/00106).

